# CPT/HCPCS Code Recommendation from Clinical Notes: A Comparative Evaluation of AI Methods

**DOI:** 10.64898/2026.08.29.26361731

**Authors:** Qingyuan Song, Congning Ni, Weixin Liu, Yike Li, Bradley A. Malin, Zhijun Yin

**Author notes:** {;,;}. {, }.

## Abstract

Automatic coding from clinical notes has been studied extensively for International Classification of Diseases (ICD) codes, yet broad Current Procedural Terminology (CPT) and Healthcare Common Procedure Coding System (HCPCS) recommendation remains comparatively underexplored. Existing studies often focus on one specialty, a limited code vocabulary, or a single model family, leaving it unclear how different artificial intelligence (AI) paradigms perform under a common, clinically meaningful evaluation. We formulate CPT and HCPCS coding as an AI-assisted recommendation task in which a physician or professional coder reviews a short, ranked list of candidate codes supported by the clinical note. Using operative notes from Vanderbilt University Medical Center (VUMC) and discharge summaries from Medical Information Mart for Intensive Care IV (MIMIC-IV), we compare lexical retrieval, Clinical-Longformer, GPT-5.6-Sol, MedGemma-27B, and an inspectable agentic-style retrieve-and-verify system under a controlled review budget. Micro-averaged recall within a fixed number of recommendations measures whether reference codes reach the reviewable list; micro-F1 is reported only where reference labels are sufficiently complete. Zero-shot GPT-5.6-Sol achieves the highest recall within five and ten candidates: 0.717 and 0.800 on VUMC and lower-bound values of 0.689 and 0.738 on MIMIC-IV. The retrieve-and-verify system reaches 0.695 and 0.784 on VUMC and lower-bound values of 0.575 and 0.657 on MIMIC-IV, with a candidate-linked evidence window attached to each retained recommendation. Diagnostic analyses reveal distinct failure sources, including output-length underfilling, confusion among closely related codes, out-of-knowledge-base generation, and incomplete evidence support. These findings establish a systematic evaluation framework for procedure-code recommendation and identify practical requirements for future systems that are accurate, review-efficient, and grounded in clinical evidence.

## I. Introduction

Current Procedural Terminology (CPT) and the Healthcare Common Procedure Coding System (HCPCS) provide the standard vocabularies used to report procedures, services, products, and supplies in U.S. health care [1], [2]. These codes support claims processing and reimbursement and also contribute to utilization measurement, quality reporting, and health-services research. Accurate coding therefore matters not only for individual claims but also for the administrative and analytic data used to characterize delivered care. Yet assigning CPT/HCPCS codes remains challenging since the decisive procedural details are often embedded in long and heterogeneous clinical notes.

Automatic coding from clinical notes has been studied extensively, but most prior work has focused on International Classification of Diseases (ICD) diagnosis codes. Representative approaches include code-specific attention models [3], hierarchical and label-attention architectures [4], long-document encoders [5], pretrained language models with label-wise attention [6], synonym-aware label matching [7], calibrated label embeddings [8], and retrieval and reranking [9]. Procedure coding has received comparatively less attention. Existing CPT studies have more often been restricted to particular specialties or limited code vocabularies, including pathology reports [10], common musculoskeletal procedures [11], selected cervical-spine operations [12], and feasibility studies with large language models on clinic notes [13]. More recently, CPTCoder and the associated MIMIC-CPT resource have provided a broader benchmark [14], but evidence remains fragmented across note types, code vocabularies, and model families.

Procedure-code recommendation is not simply diagnosis coding with a different label vocabulary. Fine-grained CPT/HCPCS selection often requires reconstructing what was actually performed from procedural details distributed across the clinical narrative. Relevant evidence may include the anatomic site and laterality, procedural approach, device or material used, extent of work, diagnostic versus therapeutic intent, and additional services performed during the same encounter. Closely related codes may share most of their descriptions while differing in only one such detail. Therefore, procedure code recommendation must retrieve plausible candidates and discriminate among closely related codes using evidence that may span multiple sentences.

It remains unclear how different modeling paradigms compare for broad CPT/HCPCS recommendation under a common protocol. They consume notes, represent the code space, and determine output length differently. A sparse retriever may naturally return a long ranked list, whereas a generative model may stop after one or two codes. Comparing their recall at a nominal cutoff therefore obscures differences in review burden. Note inputs can differ across systems, and label completeness varies across data sources, making precision and F1 difficult to compare across corpora. A credible comparison must document input and label policies, control the review budget, distinguish ranking from set prediction, account for label completeness, and quantify uncertainty in observed performance.

We consequently frame the task as AI-assisted code recommendation rather than autonomous billing. A physician or professional coder reviews a short ranked list produced by a system and decides which codes the note supports. Under this intended use, the central question is not whether a model independently reconstructs the complete final claim, but whether the relevant codes are surfaced within a reviewable recommendation set. This motivates micro-averaged Recall@*N* as the primary metric because it measures the fraction of reference codes recovered within a review budget of *N* candidates. Micro-F1 evaluates a separate and set-prediction task in which the number of predicted codes is not fixed, because F1 penalizes both missed codes and additional predictions. And we report it only when the available reference-code sets are sufficiently complete.

Guided by this framing, we conduct a two-corpus evaluation of broad CPT/HCPCS recommendation using private operative notes from Vanderbilt University Medical Center (VUMC) and credentialed-access discharge summaries from MIMIC-IV. Following the general AI-supported coding practice, we compare lexical retrieval, long-document classification, zero-shot generation with a proprietary large language model (LLM), retrieval-augmented generation (RAG), and supervised fine-tuning of an open-weight LLM under documented input policies and a common review budget. Because CPT/HCPCS coding involves distinguishing among closely related candidate codes and determining whether the documentation provides sufficient evidence for each code, we further design and evaluate an inspectable agentic-style retrieve-and-verify system that decomposes coding into candidate generation and evidence-based verification before presenting a shortlist for human review. Our work makes three contributions:

1. **A controlled two-corpus evaluation of procedure-code recommendation**. We compare six approaches spanning retrieval, classification, zero-shot generation, RAG, and supervised fine-tuning on VUMC operative notes and MIMIC-IV discharge summaries under documented input policies and a review-budget protocol.
2. **A compact, inspectable retrieve-and-verify system**. We introduce a 110-million-parameter modular system that combines note-type-aware sectionization, complementary candidate retrieval, and evidence-conditioned verification, while exposing supporting note evidence and intermediate retrieval behavior for each recommendation.
3. **A diagnostic analysis of performance, failure modes, and review burden**. Beyond Recall@*N* , we analyze review burden, uncertainty, retrieval ceilings, code-frequency effects, substitution errors, out-of-knowledge-base generation, and evidence support to distinguish genuine model improvements from gains driven by larger candidate lists.

Together, these findings establish an evaluation foundation for developing future procedure-code recommendation systems that move beyond accuracy alone toward reliable, review-efficient, and evidence-grounded coding support.

## II. Related Work

### A. Neural Clinical Coding

Most automated clinical-coding research treats ICD assignment as extreme multi-label classification. Convolutional Attention for Multi-Label classification (CAML) introduced code-specific attention [3], and the Label Attention Model (LAAT) combined label-specific attention with hierarchical learning for rare codes [4]. Clinical-Longformer extended usable context for long clinical notes [5]. Pretrained Language Model for ICD Coding (PLM-ICD) combined pretrained encoders with segment pooling and label-wise attention [6]. Other work enriched labels with synonyms [7], calibrated attention over diverse label embeddings [8], and retrieved and reranked candidate codes [9]. Together, these studies showed the technical progression of architectures for note-to-code modeling, but they primarily targeted ICD rather than mixed CPT/HCPCS procedure vocabularies.

### B. CPT and HCPCS Procedure Coding

The smaller CPT literature demonstrated feasibility but usually limited the vocabulary or note type. Existing studies predicted five primary and 38 ancillary CPT codes from pathology reports [10], the 100 most common musculoskeletal CPT codes [11], or selected cervical-spine procedures rather than the full procedure vocabulary [12]. GPT-4 evaluation of CPT and HCPCS generation from clinic notes reported a 28.9% mean true-positive rate, illustrating the difficulty of unconstrained generation [13]. CPTCoder added constrained decoding, per-code confidence, a MIMIC-CPT benchmark, and an external hospital evaluation [14]. Our work complements these studies by comparing distinct method families under a common review-budget metric, testing two note genres, documenting input differences, and measuring the shortlist that a human actually reviews.

### C. Retrieval, Reranking, and Generation

Large label spaces can also be reduced before classification. Multi-stage systems retrieve and rerank medical codes [9], and biomedical entity encoders such as SapBERT match code descriptions to clinical text [15]. Retrieval-augmented generation (RAG) combines retrieved non-parametric context with a generative model [16], while low-rank adaptation (LoRA) makes supervised adaptation of large models more tractable [17]. Our evaluation represents all three choices: code retrieval, exemplar-based RAG, and supervised fine-tuning (SFT) of MedGemma-27B [18]. The agentic-style retrieve-and-verify system instead retrieves code candidates, pairs each candidate with note evidence, and learns a discriminative verifier. This modularity makes retrieval and ranking failures separately inspectable.

## III. Evaluation Design

### A. Task Definition

Let *x*_*n*_ denote the clinical note for encounter *n*, let *C* denote the canonical vocabulary of modifier-free Current Procedural Terminology (CPT) and Healthcare Common Procedure Coding System (HCPCS) base codes, and let

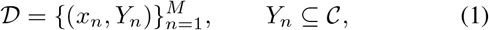

be a labeled corpus of *M* encounters, where *Y*_*n*_⊆ *C* is the reference-code set. An AI system is required to return a ranked shortlist *r*_*n*_ = (*c*_*n*,1_, *c*_*n*,2_, …) and, when set prediction is evaluated, a decoded set *Ŷ*_*n*_. Modifiers and service units are outside the prediction target because their assignment primarily depends on external administrative, contractual, and payer-specific information rather than on the clinical note alone. Restricting the target to base codes also avoids fragmenting the label space into sparse modifier and unit combinations. Ranked recommendation is primary: Recall@*N* measures how much of *Y*_*n*_ appears within the first *N* positions of *r*_*n*_. Set micro-F1 evaluates *Ŷ*_*n*_ only when the reference-code set is considered sufficiently complete. Historical base codes remain valid on their documented procedure dates, and date-aware crosswalks are applied when possible.

### B. Corpora

#### 1) VUMC Operative Notes

The VUMC corpus contains raw operative notes linked to billed CPT and HCPCS base codes. We draw an analytic sample after preprocessing, and the chronological split contains 19,627 training notes from 2017–2023, 1,975 validation notes from 2024, and 1,985 test notes from 2025. These splits contain 35,768, 3,849, and 3,900 reference-code instances, respectively. Raw clinical text can contain literal billing-code strings. A leakage scrubber removes code-shaped strings before indexing or inference. The scrubber affected approximately 42% of linked reference-code instances, and post-scrub validation found no remaining verbatim reference-code leakage. All label-derived resources use the training split only.

#### 2) MIMIC-IV Discharge Summaries

The MIMIC-IV corpus joins MIMIC-IV v3.1 procedure billing records to MIMIC-IV-Note v2.2 discharge summaries by hospital admission [19]– [21]. MIMIC-IV-Note contains 331,794 discharge summaries in its source release. The final cohort contains 14,813 surgery-coded admissions with a linked discharge summary in a patient-disjoint 70/10/20 split. It contains 10,361 training, 1,481 validation, and 2,971 test notes, with 16,107, 2,274, and 4,698 recorded procedure-code instances, respectively. We select encounters containing at least one surgical CPT code and remove nonprocedural codes that cannot be inferred from the narrative. This filter raises the surgical share of recorded reference-code instances from 61.60% to 99.48%. Only 43.81% of admissions with a discharge summary have any procedure billing record, so MIMIC-IV reference-code sets are treated as partially recorded. Table I summarizes the corpus characteristics and evaluation contract.

**TABLE I.** Corpus characteristics and evaluation contract.

| Characteristic | VUMC | MIMIC-IV |
| --- | --- | --- |
| Note type | Operative note | Discharge summary |
| Access | Private | Credentialed access |
| Split policy | Chronological | Patient-disjoint 70/10/20 |
| Train / validation / test notes | 19,627 / 1,975 / 1,985 | 10,361 / 1,481 / 2,971 |
| Reference-code instances, train / validation / test | 35,768 / 3,849 / 3,900 | 16,107 / 2,274 / 4,698 |
| Mean codes per encounter | 1.845 | 1.558 |
| Surgical share of reference-code instances | 93.78% | 99.48% after procedural filter |
| abel completeness | Reference-complete for this study | Partially recorded; 43.81% have any HCPCS record |

### C. Code Knowledge Base

We construct a knowledge base (KB) from a fixed 2026 CPT/HCPCS code-reference snapshot from Codify by AAPC [22]. It contains 19,780 CPT/HCPCS rows, including 18,923 codes active in 2026. Each code is associated with its official description and a hierarchy of up to six ranges. For example, CPT 93000, standard electrocardiogram with interpretation and report, has the three-level hierarchy Medicine Services and Procedures; Cardiovascular Procedures; and Cardiography Procedures. A separate historical add-on contains 13,835 new, revised, or deleted code events across 12,220 codes, including 2,736 resolved deleted-to-substitute mappings. Combining these resources gives 27,661 codes in total.

### D. Compared Systems

- **M1 — BM25 neighbor voting**. BM25 [23] retrieves the 25 most similar training notes. Codes in neighbor reference sets receive similarity-weighted votes, and a validation-tuned threshold produces the final set.
- **M2 — Clinical-Longformer with label attention**. Clinical-Longformer encodes up to 4,096 tokens [5], and a PLM-ICD-style label-attention head learns a separate representation for each code [6]. Codes with training frequency below five are outside its classifier vocabulary.
- **M3 — Zero-shot GPT-5.6-Sol**. The large language model (LLM) reads the clinical note without examples or task-specific training and returns a JavaScript Object Notation (JSON). We evaluate two output settings. The *10-code setting* requests ten ranked codes using prompt instruction and supplies the ranking prediction; the *unrestricted setting* requests no fixed length and supplies the set prediction.
- **M4 — GPT-5.6-Sol exemplar RAG**. BM25 retrieves three diverse training-note examples. Their reference codes form a candidate pool from which GPT-5.6-Sol selects. The system uses the 10-code setting for both its ranking and generated set.
- **M5 — LoRA SFT MedGemma-27B**. A separate MedGemma-27B adapter is fitted on each corpus’s training split with low-rank adaptation (LoRA): rank 16, scaling 32, dropout 0.05, two epochs, and completion-only loss. It uses the zero-shot GPT-5.6-Sol prompt format and parser. As with zero-shot GPT-5.6-Sol, we evaluate a 10-code setting for ranking output and an unrestricted setting for set output.
- **M6 — Agentic-style retrieve-and-verify system**. A note-type-aware sectionizer can retain procedure-oriented sections or pass the whole note. The resulting text forms overlapping sentence-aligned windows, retrieves candidates from three sources, and scores each candidate against its strongest window. Its sectionizer, retriever, verifier, and learning objective are defined below.

### E. Input and Label Processing

The common target input is a scrubbed prefix of at most 4,096 `cl100k_base` tokens. The Clinical-Longformer, GPT-5.6-Sol, and MedGemma-27B systems consume this prefix, while BM25 neighbor voting is uncapped. The retrieve-and-verify system supports a whole-note view and a note-type-specific selected-section view, each converted into at most 32 evidence windows.

Codes are normalized to uppercase, modifier-free base forms. For generative systems, every code-shaped string is tallied before output validation, a string absent from the current and historical knowledge base (KB) is counted as an out-of-KB generation. Within each corpus, head codes cover the first 80% of cumulative training occurrences, torso codes the next 15%, tail codes the final 5%, and unseen codes have no training occurrence. Document section taxonomies are defined separately for operative notes and discharge summaries.

### F. Agentic-Style Retrieve-and-Verify System

Fig. 1 summarizes the system. A frozen note-type configuration first maps variable headers to canonical sections. A view policy then retains either the procedure-oriented subset or the whole sectionized note, after which the text becomes sentence-aligned evidence windows. Lexical, dense, and similar-note retrieval form a candidate pool, and a cross-encoder scores each candidate against its strongest evidence window. The output is a ranked shortlist with one evidence window per recommendation. The LLM induces header patterns offline but is not called during note-time inference.

**Fig. 1.**
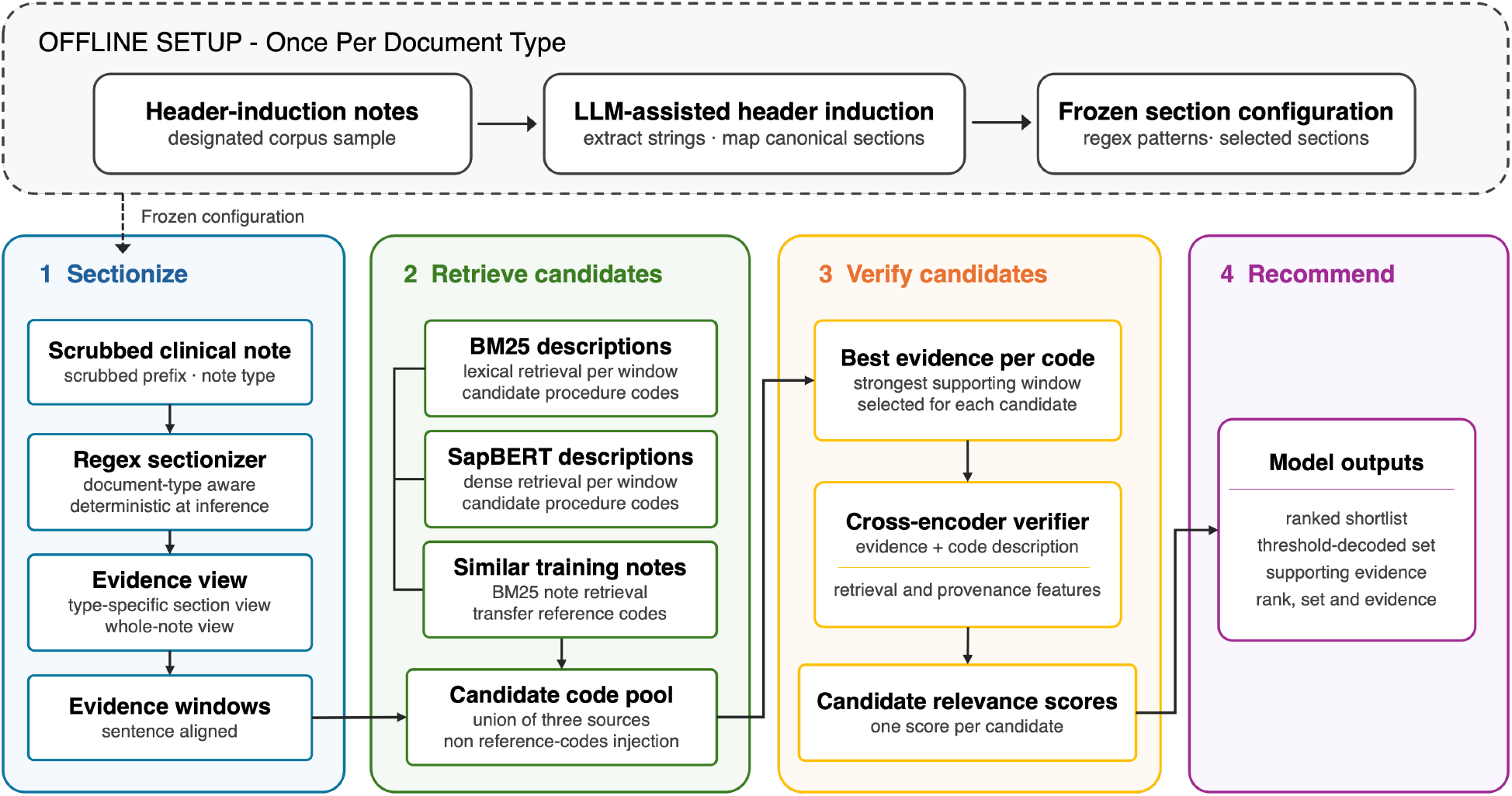
Overview of the agentic-style retrieve-and-verify system. Dashed arrows denote offline header induction; solid arrows denote note-time prediction.

We use agentic-style to describe this explicit sequence of specialized stages–sectionize, retrieve, verify, and recommend–rather than an autonomous coding agent. The sectionizer serves as the system’s attention filter by suppressing historical or administrative context that resembles the current procedure but does not document what was performed.

#### 1) Note-type-aware sectionization

Clinical-note headers vary across institutions, templates, and note types. For note type 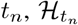 is a canonical section taxonomy and 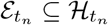 is its procedure-oriented subset. Their names and sizes are corpus-specific.

The sectionizer is induced once and then frozen. On a designated sample, an LLM extracts header strings verbatim and maps them to canonical sections; recurring forms become whitespace-tolerant regular expressions. Note-time inference uses deterministic matching only.

Let *S*_*h*_(*x*_*n*_) be the text assigned to section *h*, and let *G* create sentence-aligned evidence windows. The retained evidence-window set is

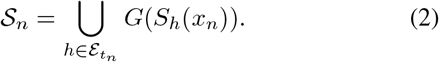

Each evidence window contains at most 180 words with 60-word overlap; at most 32 are retained with their canonical section tags. For the whole-note view, the same windowing function is applied to the complete sectionized note rather than only to 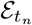.

#### 2) Multi-source candidate generation

Let *d*_*c*_ be the knowledge-base description of code *c*, and let *q* be the SapBERT encoder. BM25 and dense search each retrieve 25 descriptions per evidence window; a third source retrieves 25 training notes from the concatenated retained sections and transfers their reference codes. Let Lex_25_(*s*) and Den_25_(*s*) denote the two top-25 code sets, and let Nbr_25_(*S*_*n*_) denote the indices of the 25 retrieved training notes. The candidate pool is

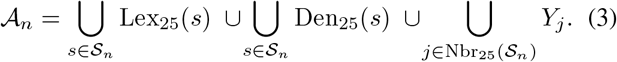

Training-note self-retrieval is excluded. Duplicate codes collapse during the union, while source membership, scores, and ranks are retained. The target set *Y*_*n*_ is never injected into *A*_*n*_.

#### 3) Evidence-conditioned candidate scoring

Each candidate is paired with the window most similar to its description. A cross-encoder jointly reads that evidence and the code description, then combines its pooled representation with a 14-dimensional feature vector *ϕ*_*n*,*c*_ containing source presence, ranks, scores, neighbor frequency, window similarity and position, source count, code-system identity, and description length:

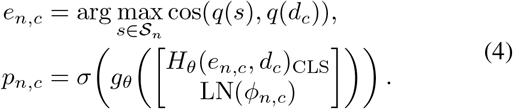

Here *H*_*θ*_ is the SapBERT-initialized cross-encoder, *g*_*θ*_ is a two-layer Gaussian error linear unit (GELU) head with dropout, LN is layer normalization, and *p*_*n*,*c*_ is the predicted probability that code *c* belongs to note *n*. The square brackets denote feature concatenation.

#### 4) Training and inference

Training uses every positive code present in the candidate pool and samples up to ten negative candidates per positive. If a note has no reachable positive, it contributes at most 20 negatives. Let *P* be the resulting training pairs and *y*_*n*,*c*_ = ⊮[*c* ∈ *Y*_*n*_]. The verifier minimizes binary cross-entropy (BCE)

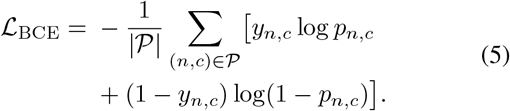

Reference codes absent from the candidate pool are not added as training examples and remain false negatives in end-to-end evaluation.

At inference, candidates are sorted by *p*_*n*,*c*_ to form *r*_*n*_. Recall@*N* uses this unthresholded top-*N* ranking shortlist. Separately, a validation-selected threshold *τ*^∗^ defines the system’s decoded code set *Ŷ*_*n*_ for the supporting micro-F1 and set-error analysis.

### G. Evaluation Metrics

The primary evaluation metric is micro-averaged Recall@*N* , where *N* is the review budget:

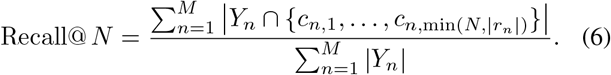

Recall@5 is primary and Recall@10 is secondary. Five candidates provide a practical review budget relative to the 1.845 VUMC and 1.558 recorded MIMIC-IV codes per encounter; Recall@10 tests a longer list. Shown@*N* is the mean number of codes actually displayed within that budget. Family mean reciprocal rank (FamilyMRR) diagnoses sibling discrimination. For reference code *g*, fam(*g*) is its deepest defined KB range, *s*_*n*_(*g*) = (*c*_*n*,*j*_ : fam(*c*_*n*,*j*_) = fam(*g*)) is the family-matched subsequence of *r*_*n*_, and pos_*n*_(*g*) is the one-based position of *g* in that subsequence. Let *Q* = {(*n, g*) : *g* ∈ *Y*_*n*_, fam(*g*) is defined, *g* ∈ *r*_*n*_}. Then

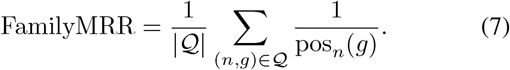

Unlike standard mean reciprocal rank, unrelated candidates do not affect the position, and candidate-pool misses are excluded rather than scored as zero. Short or sibling-sparse candidate lists can inflate the metric, so we use FamilyMRR only as a within-system diagnostic.

We use *ranking output* for Recall@*N* and FamilyMRR and *set output* for micro-F1 and false-positive analyses. For zero-shot GPT-5.6-Sol and MedGemma-27B, ranking metrics use the 10-code setting and set metrics use the unrestricted setting. For the retrieve-and-verify system, ranking metrics use unthresholded top-*N* scores and set metrics use its threshold-decoded set. BM25 neighbor voting and Clinical-Longformer use validation-fixed thresholds, whereas exemplar RAG uses its generated 10-code-setting output. Candidate-pool ceiling separates retrieval misses from verification misses.

MIMIC-IV’s `hcpcsevents` table is partially recorded. A plausible unrecorded prediction would be counted as a false positive. There will be a larger penalty for systems that emit more codes. We therefore suppress precision-derived metrics, including micro-F1 and false-positive rates on MIMIC-IV and report Recall@*N* and FamilyMRR as lower bounds. Shown@*N* depends only on predictions and remains directly interpretable.

Evidence support and coding validity are only diagnostics rather than substitutes for reference-code recovery. MIMIC Documents Annotated with Code Evidence (MDACE) demonstrates the value of coder-annotated evidence spans [24], but neither corpus supplies such spans. We therefore use an LLM judge to rate a stratified sample of 200 retrieve-and-verify evidence windows as fully, partially, or not supporting the associated code. Two human judges independently rate the same 50 sampled note–code evidence pairs on this three-point ordinal scale.

### H. Experiment Setup

BM25 neighbor voting uses BM25 over training notes, and the long-document classifier uses Clinical-Longformer [25]. The zero-shot and exemplar-RAG systems use the private GPT-5.6-Sol model. The supervised open-weight system adapts MedGemma-27B [26], while the retrieve-and-verify system uses SapBERT [27]. Unlike the general-purpose LLMs, the proposed system is a task-specific cross-encoder with 110 million parameters. MedGemma-27B has approximately 245 times as many parameters. GPT-5.6-Sol’s parameter count is not disclosed.

The VUMC configuration maps 529 observed header patterns to 19 canonical operative-note sections and retains six: Procedure(s) Performed, Detailed Description, Findings, Specimens Removed, Implants & Equipment, and Indications for Surgery. Header forms must occur at least twice, and at most 100 patterns are retained per section. The configuration was induced once from 2,000 sampled notes and then applied unchanged to all VUMC splits. MIMIC-IV uses a separate 26-category discharge-summary taxonomy and an 801-pattern bank induced from its sampled notes. Its three-section evidence view contains Procedures, Hospital Course, and Results. The retrieve-and-verify system is evaluated in whole-note and selected-section views on both corpora: six selected sections on VUMC and three on MIMIC-IV. Each MedGemma-27B adapter is fitted separately on its corpus; both output settings use that same adapter.

## IV. Results

### A. VUMC Test Comparison

Table II reports the VUMC test results. For ranking, zero-shot GPT-5.6-Sol in the 10-code setting leads Recall@5/Recall@10 at 0.717/0.800. The selected-section retrieve-and-verify system follows at 0.695/0.784, while its whole-note view reaches 0.675/0.769. GPT-5.6-Sol exemplar RAG reaches 0.590/0.601, and LoRA SFT MedGemma-27B reaches 0.579/0.598. Exemplar RAG reports a FamilyMRR of 0.935, which must be interpreted only as a within-system diagnostic and alongside its short output. For set prediction, zero-shot GPT-5.6-Sol in the unrestricted setting leads micro-F1 at 0.616, followed by unrestricted LoRA SFT MedGemma-27B at 0.558, the threshold-decoded selected-section system at 0.556, and the whole-note system at 0.542.

**TABLE II.** VUMC TEST RESULTS. M3–M5 RANKING METRICS USE THE 10-CODE SETTING; M3 AND M5 MICRO-F1 VALUES USE UNRESTRICTED GENERATION.

| ID | System | Recall@5 [95% CI] | Recall@10 | Shown@5/10 | FamilyMRR | Set micro-F1 [95% CI] |
| --- | --- | --- | --- | --- | --- | --- |
| M1 | BM25 neighbor voting | 0.593 [0.574, 0.611] | 0.680 | 4.54 / 8.11 | 0.839 | 0.482 [0.464, 0.498] |
| M2 | Clinical-Longformer | 0.520 [0.501, 0.538] | 0.603 | 5.00 / 10.00 | 0.774 | 0.365 [0.351, 0.379] |
| M3 | Zero-shot GPT-5.6-Sol | **0.717 [0.700, 0.733]** | **0.800** | 4.98 / 9.93 | 0.901 | **0.616 [0.599, 0.633]** |
| M4 | GPT-5.6-Sol exemplar RAG | 0.590 [0.571, 0.607] | 0.601 | 3.73 / 4.56 | 0.935 | 0.362 [0.349, 0.373] |
| M5 | LoRA SFT MedGemma-27B | 0.579 [0.562, 0.596] | 0.598 | 2.32 / 3.03 | 0.929 | 0.558 [0.531, 0.581] |
| M6 | Retrieve-and-verify, six sections | 0.695 [0.680, 0.712] | 0.784 | **5.00 / 10.00** | 0.817 | 0.556 [0.540, 0.571] |
| M6 | Retrieve-and-verify, whole note | 0.675 [0.658, 0.692] | 0.769 | **5.00 / 10.00** | 0.823 | 0.542 [0.527, 0.558] |

Fig. 2(a) traces VUMC Recall@*N* across review budgets. Zero-shot GPT-5.6-Sol nearly fills its 10-code request and remains strongest as the budget grows. Exemplar RAG plateaus with 4.56 codes shown at a budget of ten because it selects only from its retrieved candidate pool. MedGemma-27B likewise plateaus with 3.03 codes shown at ten, leaving much of the available review budget unused.

**Fig. 2.**
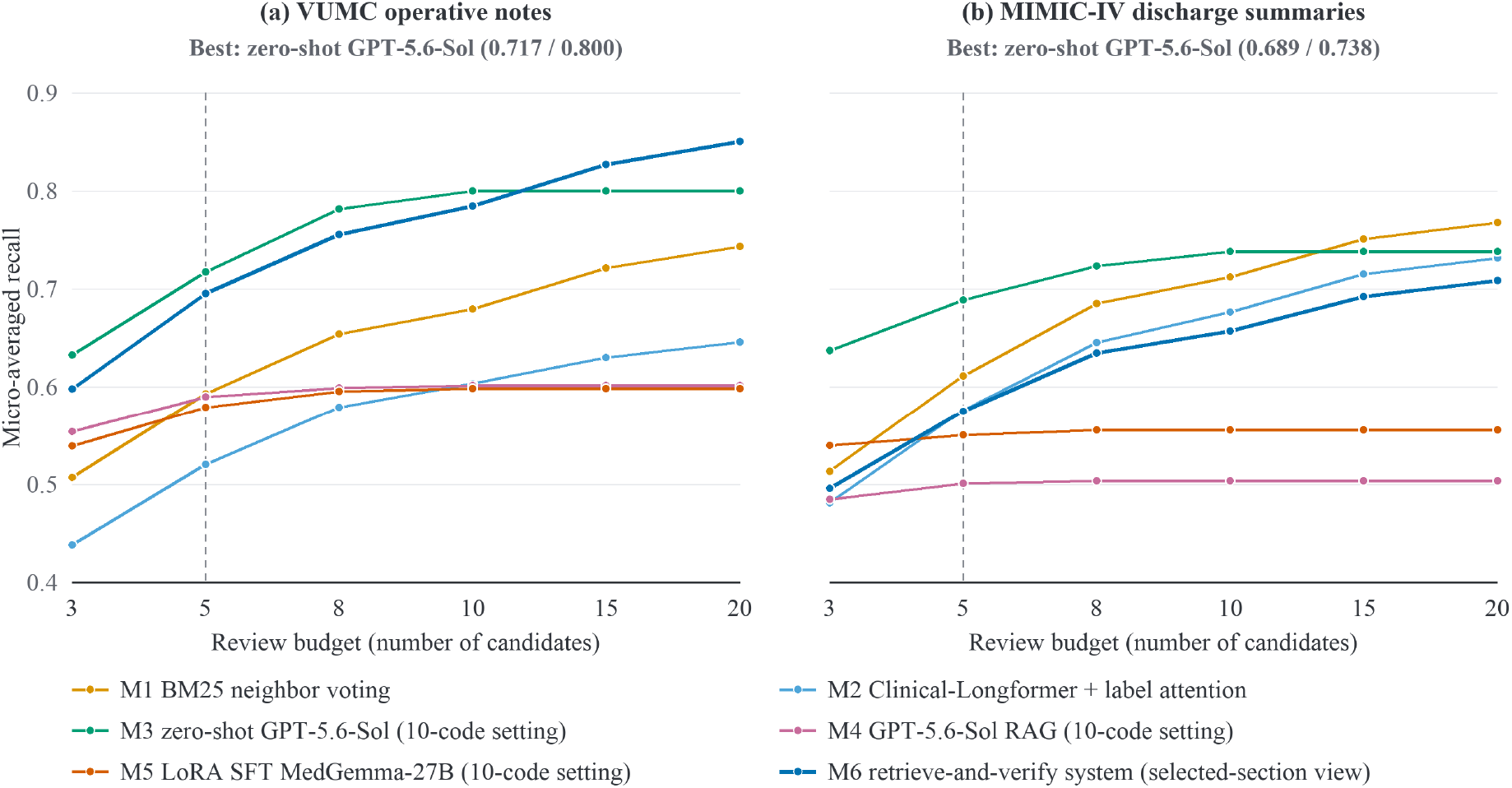
Recall@*N* by review budget on VUMC and MIMIC-IV. Zero-shot GPT-5.6-Sol, exemplar RAG, and LoRA SFT MedGemma-27B use the 10-code setting; the retrieve-and-verify curves use the selected-section view. MIMIC-IV values are lower bounds because its procedure-code records are incomplete.

### B. MIMIC-IV Replication

Table III reports the MIMIC-IV results. Recall@*N* and FamilyMRR values are lower bounds because the recorded billing records are incomplete, no set micro-F1 is reported. Zero-shot GPT-5.6-Sol leads Recall@5/Recall@10 at 0.689/0.738, followed by BM25 neighbor voting at 0.611/0.712. The whole-note and selected-section views of retrieve-and-verify system reach 0.578/0.662 and 0.575/0.657, respectively. Clinical-Longformer reaches 0.576/0.676. The separately fine-tuned MedGemma-27B reaches 0.550/0.556, and exemplar RAG is last at 0.501/0.503. MedGemma-27B reports a FamilyMRR of 0.903, but this within-system diagnostic must be read alongside its short output.

**TABLE III.** MIMIC-IV PATIENT-DISJOINT TEST RESULTS.

| ID | System | Recall@5 | Recall@10 | Shown@5/@10 | FamilyMRR |
| --- | --- | --- | --- | --- | --- |
| M1 | BM25 neighbor voting | 0.611 [0.595, 0.627] | 0.712 | 4.85 / 9.15 | 0.763 |
| M2 | Clinical-Longformer | 0.576 [0.560, 0.592] | 0.676 | 5.00 / 10.00 | 0.713 |
| M3 | Zero-shot GPT-5.6-Sol (10-code setting) | **0.689 [0.676, 0.702]** | **0.738** | 4.98 / 9.59 | 0.858 |
| M4 | GPT-5.6-Sol exemplar RAG (10-code setting) | 0.501 [0.485, 0.517] | 0.503 | 3.35 / 3.67 | 0.902 |
| M5 | LoRA SFT MedGemma-27B (10-code setting) | 0.550 [0.536, 0.566] | 0.556 | 1.88 / 2.06 | 0.903 |
| M6 | Retrieve-and-verify system, whole note | 0.578 [0.562, 0.594] | 0.662 | **5.00 / 10.00** | 0.741 |
| M6 | Retrieve-and-verify system, three sections | 0.575 [0.560, 0.591] | 0.657 | **5.00 / 10.00** | 0.739 |

Fig. 2(b) shows the MIMIC-IV curves separately. Zero-shot GPT-5.6-Sol remains strongest through a budget of ten and shows 9.59 codes at that cutoff. Exemplar RAG shows 3.67 codes and MedGemma-27B only 2.06, causing both curves to flatten early. Recall@*N* should therefore be interpreted with Shown@*N* : *N* is the maximum review budget, not a guaranteed list length.

### C. Diagnostic and Error Analysis

#### 1) Frequency Strata Expose Different Failure Mechanisms

Table IV compares Recall@5 ranking outputs by training-frequency stratum. On VUMC, the selected-section system improves every stratum over its whole-note view and leads head-code recall at 0.800. Zero-shot GPT-5.6-Sol leads torso-, tail-, and unseen-codes at 0.651, 0.597, and 0.373. BM25 neighbor voting, Clinical-Longformer, and exemplar RAG recover no unseen VUMC code because their prediction spaces derive from training labels. On MIMIC-IV, BM25 neighbor voting leads head-code recall at 0.759, while zero-shot GPT-5.6-Sol leads torso-, tail-, and unseen-codes at 0.593, 0.545, and 0.418. The selected-section view is slightly lower than the whole-note view on head and torso codes and unchanged after rounding on tail and unseen codes. MedGemma-27B unseen-code recall remains only 0.013 on VUMC and 0.014 on MIMIC-IV.

**TABLE IV.** Recall@5 BY CORPUS-SPECIFIC TRAINING-FREQUENCY STRATUM.

| System | VUMC |  |  |  | MIMIC-IV |  |  |  |
| --- | --- | --- | --- | --- | --- | --- | --- | --- |
|  | Head | Torso | Tail | Unseen | Head | Torso | Tail | Unseen |
| M1 — BM25 neighbor voting | 0.738 | 0.300 | 0.036 | 0.000 | **0.759** | 0.170 | 0.028 | 0.000 |
| M2 — Clinical-Longformer | 0.699 | 0.040 | 0.000 | 0.000 | 0.738 | 0.042 | 0.000 | 0.000 |
| M3 — Zero-shot GPT-5.6-Sol (10-code setting) | 0.761 | **0.651** | **0.597** | **0.373** | 0.730 | **0.593** | **0.545** | **0.418** |
| M4 — GPT-5.6-Sol exemplar RAG (10-code setting) | 0.705 | 0.381 | 0.161 | 0.000 | 0.599 | 0.240 | 0.109 | 0.000 |
| M5 — LoRA SFT MedGemma-27B (10-code setting) | 0.721 | 0.275 | 0.077 | 0.013 | 0.652 | 0.305 | 0.081 | 0.014 |
| M6 — Retrieve-and-verify system (whole-note view) | 0.785 | 0.460 | 0.327 | 0.082 | 0.683 | 0.304 | 0.118 | 0.058 |
| M6 — Retrieve-and-verify system (selected-section view) | **0.800** | 0.486 | 0.371 | 0.139 | 0.680 | 0.299 | 0.118 | 0.058 |

#### 2) Section Selection and Input Reduction

Section selection changes both context length and ranking. On VUMC, the six-section view lowers the test-set mean from 1,457.0 to 980.6 tokens (32.7%) while improving Recall@5/Recall@10 by 0.020/0.015. On MIMIC-IV, the three-section view lowers the mean from 2,029.5 to 563.1 tokens (72.3%) while recall decreases by only 0.003/0.005. Thus, section selection substantially reduces context, but its effect on recall depends on note type. Shorter inputs should support faster inference.

#### 3) Within-Family Errors

Two codes are siblings when they share the deepest available CPT/HCPCS range but are not identical. A sibling false positive belongs to the same family as at least one reference code. It is also a substitution false positive when that reference sibling is missing. Thus, substitution false positives are a subset of sibling false positives. Table V reports set micro-F1 and decomposes false positives under these definitions. Zero-shot GPT-5.6-Sol and MedGemma-27B uses their unrestricted setting; exemplar RAG uses its 10-code setting, and the six-section system uses its threshold-decoded set.

**TABLE V.** VUMC SET-PREDICTION AND WITHIN-FAMILY ERROR ANALYSIS.

| ID | System | Micro-F1 | False positives | Sibling-FP rate | Substitution-FP rate |
| --- | --- | --- | --- | --- | --- |
| M1 | BM25 neighbor voting | 0.482 | 1,807 | 0.395 | 0.251 |
| M2 | Clinical-Longformer with label attention | 0.365 | 3,888 | 0.386 | 0.172 |
| M3 | Zero-shot GPT-5.6-Sol (unrestricted setting) | **0.616** | 1,637 | 0.433 | 0.337 |
| M4 | GPT-5.6-Sol exemplar RAG (10-code setting) | 0.362 | 6,710 | 0.272 | 0.128 |
| M5 | LoRA SFT MedGemma-27B (unrestricted setting) | 0.558 | 1,745 | 0.481 | **0.409** |
| M6 | Retrieve-and-verify system (selected-section view) | 0.556 | 1,134 | **0.548** | 0.262 |

The selected-section system has the highest sibling-FP rate (0.548), while its substitution-FP rate is 0.262. This difference indicates that many same-family errors are additional predictions made alongside a reference code rather than replacements for a missing reference sibling. These results show that distinguishing closely related codes remains difficult. LoRA SFT MedGemma-27B has the highest substitution-FP rate (0.409). Conversely, exemplar RAG’s low substitution-FP rate (0.128) must be read alongside its 6,710 false positives and low micro-F1; a low error proportion does not imply a better set predictor when the output is much larger.

#### 4) Out-of-Knowledge-Base Generations

Table VI reports code-shaped strings that the generative systems named but that were absent from the current and historical KB. In the 10-code setting, zero-shot GPT-5.6-Sol’s out-of-KB rate is 0.655% on VUMC and 4.090% on MIMIC-IV. Its unrestricted VUMC rate is 1.180%. Candidate-constrained exemplar RAG yields 0.000% on VUMC and 0.229% on MIMIC-IV. This diagnostic captures vocabulary hallucination only; an in-KB code can still be unsupported.

**TABLE VI.** Raw out-OF-KB CODE GENERATIONS BEFORE OUTPUT VALIDATION. Rate is the percentage of named code-SHAPED STRINGS ABSENT FROM THE CURRENT AND HISTORICAL KB.

| Corpus | System | Output policy | Constraint | Named strings | Out of KB | Rate (%) |
| --- | --- | --- | --- | --- | --- | --- |
| VUMC | Zero-shot GPT-5.6-Sol | 10-code setting | KB filter | 19,850 | 130 | 0.655 |
| VUMC | Zero-shot GPT-5.6-Sol | unrestricted setting | KB filter | 4,151 | 49 | 1.180 |
| VUMC | GPT-5.6-Sol exemplar RAG | 10-code setting | Candidate filter | 9,055 | 0 | 0.000 |
| MIMIC-IV | Zero-shot GPT-5.6-Sol | 10-code setting | KB filter | 29,710 | 1,215 | 4.090 |
| MIMIC-IV | GPT-5.6-Sol exemplar RAG | 10-code setting | Candidate filter | 10,932 | 25 | 0.229 |

#### 5) Evidence Grounding

Every retained retrieve-and-verify system prediction in the audited validation ledger has an evidence window. On 200 stratified note–code pairs, the LLM judge labels 43.0% fully supported, 30.0% partially supported, and 27.0% unsupported. Full support is more common among sampled true positives than false positives (60.7% versus 20.5%). Two human judges independently rate the same 50 pairs. Their exact agreement is 0.800, with Cohen’s *κ* = 0.654. On the 40 pairs for which the two human judges agree, exact agreement between the consensus label and the LLM judge is 0.775, with *κ* = 0.619.

#### 6) Failure-Driven Improvement Priorities

The retrieve- and-verify system exhibits different bottlenecks across the two corpora. On VUMC, the selected-section system combines strong shortlist recall with the highest sibling-FP rate, indicating a need for better within-family discrimination. On MIMIC-IV, its Recall@10 remains below zero-shot GPT-5.6-Sol and BM25 neighbor voting, while the frequency-stratum results show weak rare-code coverage. These findings motivate family-balanced retrieval, sibling hard negatives, hierarchical family-to-code scoring, and expert-supervised evidence selection.

## V. Discussion

Across both corpora. zero-shot GPT-5.6-Sol’s is the strongest shortlist generator. Its 10-code setting reaches leading Recall@5 and Recall@10 at 0.717 and 0.800 on VUMC, and lower-bound values of 0.689 and 0.738 on MIMIC-IV. Neither Recall@10 value reaches complete reference-code coverage, so even the leading shortlist remains incomplete.

Set-prediction results show why ranked recommendations should not be treated as automatic code assignments. On reference-complete VUMC, unrestricted zero-shot GPT-5.6-Sol reaches the highest micro-F1 at 0.616. Unrestricted MedGemma-27B and the selected-section retrieve-and-verify system reach 0.558 and 0.556. These values leave substantial false-positive and false-negative error and do not support automatic code assignment. The results consequently support presenting candidates for human review rather than assigning codes automatically. Better calibration, abstention, and expert-supervised verification are needed before a predicted set could be used more directly.

Output length also matters. Shown@*N* reveals whether a system fills that budget. Candidate-constrained exemplar RAG plateaus at 4.56 codes on VUMC and 3.67 on MIMIC-IV. MedGemma-27B likewise shows only 3.03 and 2.06 codes at a budget of ten, affected by learning short target cardinality during supervised generation. A larger budget *N* cannot recover candidates absent from its retrieved-and-selected list. So this underfilling limits the usable depth of both shortlists and helps explain why their recall curves flatten. Future training could use distinct targets for set prediction and ranking, with siblings treated as alternatives and supervise larger cardinality as well as code order.

Frequency strata expose a shared weakness of supervised adaptation. MedGemma-27B Recall@5 declines from 0.721 on VUMC head codes to 0.275, 0.077, and 0.013 on torso-, tail-, and unseen-codes; on MIMIC-IV, it declines from 0.652 to 0.305, 0.081, and 0.014. These within-corpus declines show that the adapters favor frequent labels, motivating code-frequency-stratified sampling, rare-code oversampling, and family-aware hard negatives. Zero-shot GPT-5.6-Sol instead leads tail and unseen recall, while BM25, Clinical-Longformer, and exemplar RAG recover no unseen VUMC codes because their prediction spaces are limited to training labels.

Section selection creates a note-type dependent accuracy– context trade-off. On VUMC, it raises Recall@5/Recall@10 from 0.675/0.769 to 0.695/0.784 while reducing mean input length by 32.7%. On MIMIC-IV, it lowers recall only from 0.578/0.662 to 0.575/0.657 while reducing input length by 72.3%. Shorter inputs should support faster inference. The small MIMIC-IV loss may arise because procedures appear across different discharge-summary sections, allowing a three-section view to omit occasional evidence.

Within-family errors identify another distinct weakness. On VUMC, the selected-section system’s 0.548 sibling-FP rate shows that plausible family matches still fail exact-code discrimination. On MIMIC-IV, its Recall@10 of 0.657 trails zero-shot GPT-5.6-Sol and BM25 neighbor voting, and its rare-code results remain weak. Improvement therefore requires stronger within-family verification on VUMC and broader rare-code retrieval and ranking on MIMIC-IV

Out-of-KB generation and evidence grounding show why inspectable outputs are not automatically reliable. Zero-shot GPT-5.6-Sol’s out-of-KB rate reaches 4.1% on MIMIC-IV, and 27.0% of sampled retrieve-and-verify evidence windows are unsupported. A KB reference is needed to removes nonexistent codes, while expert-audited spans are needed to validate and improve evidence selection.

Model scale provides another practical distinction. The retrieve-and-verify system contains 110 million parameters, approximately 245-fold fewer than MedGemma-27B, and does not require an LLM during inference. This design enables local execution and reduces dependence on externally hosted model services. More broadly, it allows direct inspect on candidate retrieval, evidence selection, and verification failures can be inspected directly. GPT-5.6-Sol, in contrast, was accessed through a VUMC-approved private API. These findings suggest that model scale alone does not resolve the challenges of reliable CPT/HCPCS code recommendation.

## VI. Limitations and Ethics

This study was designated as non-human-subjects research by the Vanderbilt University Medical Center (VUMC) Institutional Review Board. The VUMC corpus contains institution-private clinical data and cannot be redistributed. Access requires appropriate institutional review, execution of a data-use agreement, and collaboration with a VUMC-affiliated investigator. All VUMC data were processed within approved local computing environments or through VUMC-approved private GPT APIs. The MIMIC-IV data are available to credentialed users through PhysioNet under its data-use requirements.

Several limitations should be considered. First, restricted access to the VUMC corpus limits independent replication, although the portable protocol and MIMIC-IV evaluation support partial external reproducibility. Second, neither corpus contains expert-annotated evidence spans linking specific note content to individual procedure codes. Although this limitation does not directly affect aggregate model evaluation, it constrains the supervision available for model development. Third, the section taxonomy and pattern bank are study-specific and lack expert boundary annotations. Fourth, the task excludes modifiers and service units. Finally, because CPT terminology is licensed and updated annually, reproducibility requires recording the Codify snapshot and vocabulary year without redistributing proprietary text.

The evaluated systems are intended to support, rather than replace, physicians and professional coders by surfacing candidate codes and associated note evidence for review. Final code assignment remains the responsibility of a qualified human reviewer. Accordingly, the present results should not be interpreted as demonstrating readiness for autonomous billing or reimbursement decisions.

## VII. Conclusion

We present a two-corpus evaluation for CPT/HCPCS procedure-code recommendation under a fixed human-review budget. Zero-shot GPT-5.6-Sol achieves the highest observed Recall@5 and Recall@10 on both corpora, yet its top ten still misses 20.0% or more reference codes. Corpus-specific fine-tuned MedGemma-27B adapters provide little rare- or unseen-code coverage and underfill the requested shortlist, showing that in-corpus supervision does not ensure adequate recall. The agentic-style retrieve-and-verify system is not the leader, but it makes candidate-retrieval, verification, and evidence-support failures easier to inspect. Section selection improves VUMC recall and preserves most MIMIC-IV recall while using fewer input tokens. A natural next step is to introduce expert-labeled evidence spans, which would provide stronger supervision for evidence selection and code verification. These findings position the system as a transparent research reference and the evaluation as a basis for future procedure-code recommendation research.

## Data Availability

Access requires appropriate institutional review, execution of a data-use agreement, and collaboration with a VUMC-affiliated investigator. The MIMIC-IV data are available to credentialed users through PhysioNet under its data-use requirements.

https://physionet.org/content/mimiciv/3.1/

https://physionet.org/content/mimic-iv-note/2.2/

## Acknowledgment

This project was supported by the Vanderbilt Innovation Catalyst Fund. We thank Donnie Sengstack, MS, Elise Russo, MPH, and Allison McCoy, PhD, for their assistance with clinical note extraction at Vanderbilt University Medical Center.

## Notes

### Competing Interest Statement

The authors have declared no competing interest.

### Author Declarations

Ethics committee/IRB of Vanderbilt University Medical Center (VUMC) waived ethical approval for this work.

